# Comparison of Structural Diagnosis and Management (SDM) approach and Myofascial Release (MFR) for improving planter heel pain, ankle range of motion and disability: A Randomized Clinical Trial

**DOI:** 10.1101/2022.08.15.22278805

**Authors:** Sapia Akter, Mohammad Shahadat Hossain, K M Amran Hossain, Zakir Uddin, Mohammad Anwar Hossain, Foisal Alom, Md. Feroz Kabir, Lori Maria Walton, Veena Raigangar

## Abstract

**Purpose:** This study compared the effect of Structural Diagnosis and Management (SDM) approach over Myofascial Release (MFR) on gastrocnemii, soleus and plantar fascia in patients with plantar heel pain.

**Subjects:** Sixty-four (n=64) subjects, aged 30-60 years, with a diagnosis of plantar heel pain, plantar fasciitis or calcaneal spur by a physician and according to ICD-10. Participants were equally allocated to MFR (n=32) and SDM (n=32) group by hospital randomization and concealed allocation.

**Methods:** In this assessor blinded randomized clinical trial, the control group performed MFR (three tissue specific stretching techniques) and the experimental group performed 2 tissue-specific interventions utilizing the Structural Diagnosis and Management (SDM) concept for 12 sessions over a 4-week period. In addition, both groups received strengthening exercises and other conventional treatments. Pain, activity limitations and disability were assessed as primary outcomes utilizing the foot function index (FFI) and range of motion (ROM) of the ankle dorsiflexors and plantar flexors were measured with a universal goniometer. Secondary outcomes were measured using the Foot Ankle Disability Index (FADI) and 10-point manual muscle testing process for the ankle dorsiflexors and plantar flexors.

**Result:** Both MFR and SDM groups exhibited significant improvements from baseline in all outcome variables, including: pain, activity level, disability, range of motion and function after the 12-week intervention period (p<.05), The SDM group showed more significant improvements than MFR for FFI pain (p=.001), FFI activity (p=.009), FFI (p= .001) and FADI (p=.002).

**Conclusion:** MFR and SDM approaches are both effective to reduce pain, improving function, ankle range of motion, and reduce disability in plantar heel pain. However, the SDM approach is significantly superior (for reducing pain, improving function and reducing disability (p<.05).

## INTRODUCTION

Plantar heel pain has a population prevalence of 7.9% and is a disabling condition of the lower limbs, manifested by pain in one or both feet [1]. This condition clinically presents with heel pain, limited ankle range of motion (ROM), reduced muscle strength of the foot and ankle fasciitis often associated with increased body mass index, and is prevalent in occupations with prolonged standing and multifactorial problems [2]. The pathophysiology includes repetitive abnormal stress, micro-tears to the plantar fascia at the insertion site causing repetitive collagen degradation, inflammation and thickening of the fascia [3] leading to localized pain in loading positions, tenderness and exaggeration of symptoms in the morning [4]. The biomechanical factors for plantar heel pain have been discussed by numerous researchers, [1-13], the most prominent theories are summarized below. The theories are based on biomechanical alteration of foot and lower limb position and mobility causing increased stress to the plantar fascia (Table 1). Moreover, Bolgla and Malone [14] described the biomechanical link with plantar fascia and windlass mechanism. Figure 1 shows an imaginary triangle drawn connecting calcaneus, midtarsal joint and metatarsal forming a “truss’. The plantar fascia (along the horizontal line in figure) acts like the “Spanish windlass”. The body weight acts in an inferior direction on both the anterior and posterior part of tibia, and midtarsal joint [15]. The ground reaction force is directed upwards in the calcaneus and metatarsal joint. The “truss” forms a stress tension that is necessary to maintain medial longitudinal arch [16]. Biomechanical abnormalities in the anterior and posterior tibial line or the structures related to the “truss” lead to increases in the stretch to plantar fascia and perifascial structures.

**Table 1:** The theories of Biomechanical abnormalities leading to increasing stress to plantar fascia and perifascial structures.

|  |  |  |
| --- | --- | --- |
| Forefoot varus causing excessive pronation during gait [5-7] | Excessive mobility or over functioning of foot beyond normal range [5,6] | Increase or decrease arch of foot causing alteration of normal mobility [12,13] |
| ⇓ | ⇓ | ⇓ |
| Excessive mobility of the foot [8] | More stress to medial joint capsules and ligamentous structures of foot [5,6] | Deviation from normal mobility required to absorb the ground reaction force through foot [12,13] |
| ⇓ | ⇓ | ⇓ |
| increased stress level to soft tissues of foot [8-10] | Adjustment of posterior tibialis by becoming lengthened and hypoactive [11] | Inability to dissipate the forces from heel strike to midstance [13] |
| ⇓ | ⇓ | ⇓ |
| Stress to plantar fascia and perifascial structures [8-10] | Pain, discomfort and increasing stress to plantar fascia and perifascial structures [5,6] | Increasing load to plantar fascia causing stress to plantar fascia and perifascial structures [12,13] |

**Figure 1:**
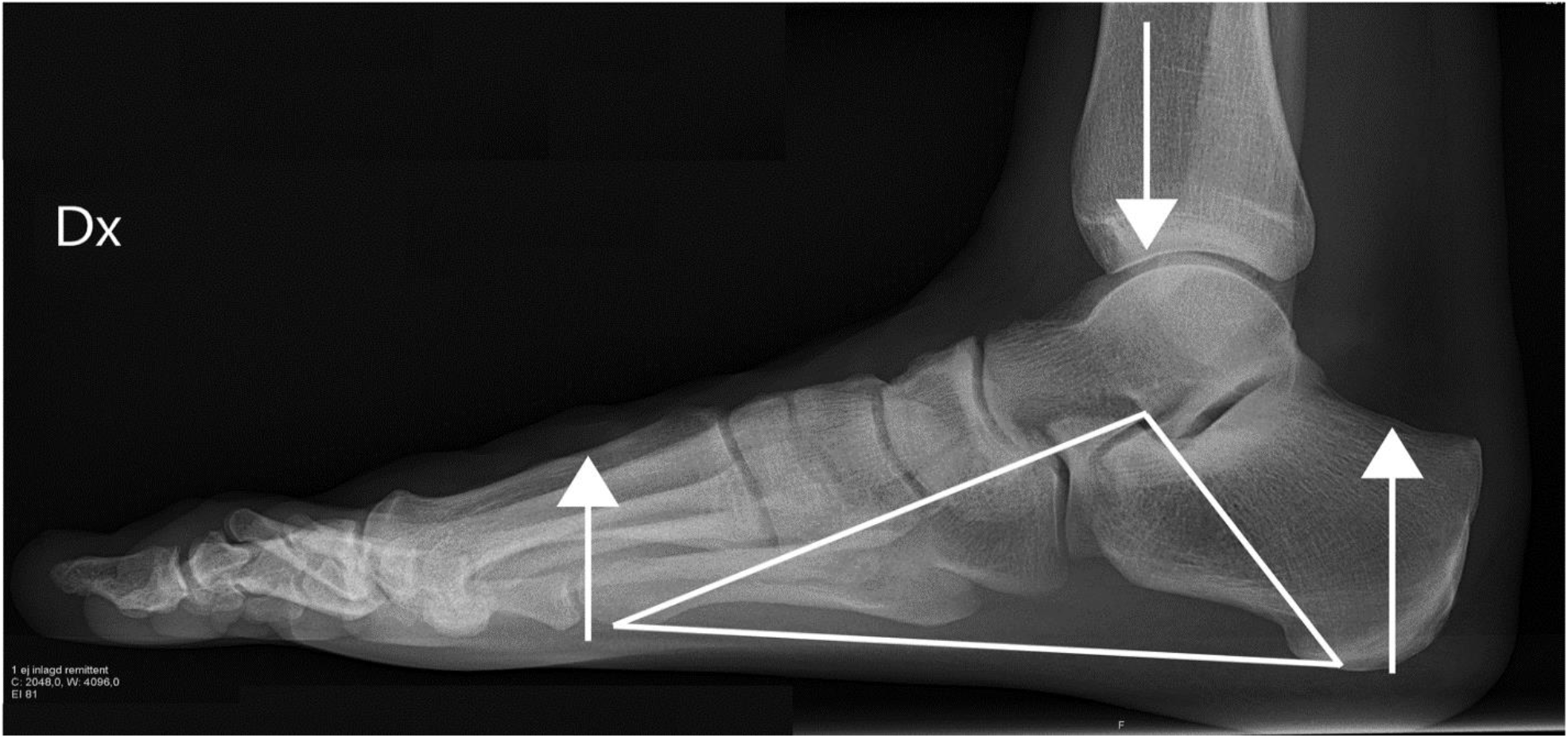
The windlass mechanism. The windlass mechanism following Bolgla and Malone, plantar fascia supports the maintenance of arch and weight distribution of stress through the feet, forefoot varus contributes to excessive pronation and higher arch during ambulation, creating more stress to the plantar musculature and fascia, and creating biomechanical abnormality in the global mobilizer of ankle. The horizontal line in the figure is the plantar fascia stress like “Spanish Windlass”

Plantar heel pain is one of the common conditions among those visiting physiotherapists. In the UK, 41.0% of patients with plantar heel pain visit a physiotherapist, regularly [17]. The treatment approach prescribed combines the use of analgesics, orthotics, splints or taping, stretching exercises, self-directed exercise, ultrasound therapy, extracorporeal shock wave therapy and corticosteroid therapy [18]. Studies suggests [19] that incorporation of a plantar fascia selective stretching program in addition to cuff stretching exercise for eight weeks can reduce pain and improve range of motion in chronic plantar fasciitis for up to two years. Some researchers suggest that stretching the plantar fascia in a non-weight bearing position is the most effective treatment protocol [20].

Myofascial release (MFR) is applied using various techniques, to reduce the tensile load of the plantar fascia at the attachment site. Common soft tissue management in MFR includes deep stripping on the plantar surface of the foot towards the calcaneus or may involve friction to the plantar fascia directed away from the calcaneus [21]. Myofascial release (MFR) over the pressure pain area of the fascia, over the calcaneus and the gastrocnemii and soleus muscles have been proven to be effective in reducing pain and improving function within 12 sessions, over 4 weeks [22]. A variety of studies suggest MFR to be superior to friction message, ultrasound therapy, stretching to the fascia in remission of pain and improvement of function [23]. The mechanism of the intervention describes stretching of the muscle components of the fascial layer, breaking cross linkages and changing viscosity of substances in the fascia [24], although the mechanism for intervention effects on ankle range, strength and function are not clear.

Structural Diagnosis and Management (SDM) [27] for plantar heel pain has been conceptualized based on the theories of biomechanical abnormalities (table 1) [5-13] and windlass mechanism [14-16]. Table 2 describes the chronology of designing the level of approaches in SDM. The treatment is directed towards the flexibility of plantar foot structures and calf muscles and improves the function of posterior leg muscles and nerves (Table 2).

**Table 2:** The theoretical concept of SDM for plantar heel pain.

|  |  |  |
| --- | --- | --- |
| Dorsiflexion stretches and dorsiflexion mobilization | Release of local trigger points of calf muscles | Facilitation of muscles and nerves of posterior leg |
| Counterforce forefoot varus causing excessive pronation during gait<br>⇓<br>Prevents excessive mobility of the foot<br>⇓<br>Reduces and normalizes stress level to soft tissues of foot<br>⇓<br>Normalizes the stress to plantar fascia and to the perifascial structures | Improves flexibility of Gastrocnemius and Soleus and promotes normal mobility of foot<br>⇓<br>enhance normal mobility required to absorb the ground reaction force through foot<br>⇓<br>Promotes normal dissipation of the forces from heel strike to midstance<br>⇓<br>Normalizes load to plantar fascia and perifascial structures | Prevents excessive mobility or over functioning of foot beyond normal range<br>⇓<br>Reduces stress to medial joint capsules and ligamentous structures of foot<br>⇓<br>Normalizes activities of posterior tibialis<br>⇓<br>Reduce pain, discomfort and decreasing stress to plantar fascia and perifascial structures |

Moreover, another study recommends [25] the understanding of the “windlass mechanism” that contributes to the biomechanical abnormalities leading to plantar heel pain. As plantar fascia supports the maintenance of arch and weight distribution of stress through the feet, forefoot varus contributes to excessive pronation and higher arch during ambulation, creating more stress to the plantar musculature and fascia, and creating biomechanical abnormality in the global mobilizer of ankle [26]. Figure 1 shows the theory behind the biomechanical correction through SDM. The key concept is to normalize the plantar fascia stress (along the horizontal line in figure) and establish a normal “Spanish Windlass”. In addition, Gastrocnemius and soleus are interconnected through fascial components connected to the plantar fascia [21]. The flexibility of this connected system of gastrocnemius, soleus and plantar fascia can direct the resultant force of the body weight downwards to both the anterior and posterior part of tibia, and mid tarsal joint [15]. This may also, enhance a biomechanical stability of the ground reaction force which acts upwards on the calcaneus and metatarsal joint, thus reduce stretch to planter fascia and perifascial structures.

SDM is a newly designed hypothetical concept and needs to be examined through a systematic process. Hence the aim of this study was to compare the effectiveness of SDM approach with MFR approach to improve pain, ankle range of motion and disability in subjects with plantar heel pain.

## SUBJECTS AND METHOD

The study was an assessor blinded, randomized clinical trial, carried out for 18 months at the Centre for the Rehabilitation of the Paralyzed, in Savar, Bangladesh. The study was approved by BHPI Institutional Review Board (IRB) and prospective trial registration was obtained from the WHO primary trial registry platform, feasibility of the study was done via a pilot RCT of 10 subjects. Consolidated Standards of Reporting Trials (CONSORT) guideline were followed for the study (Figure 2).

**Figure 2:**
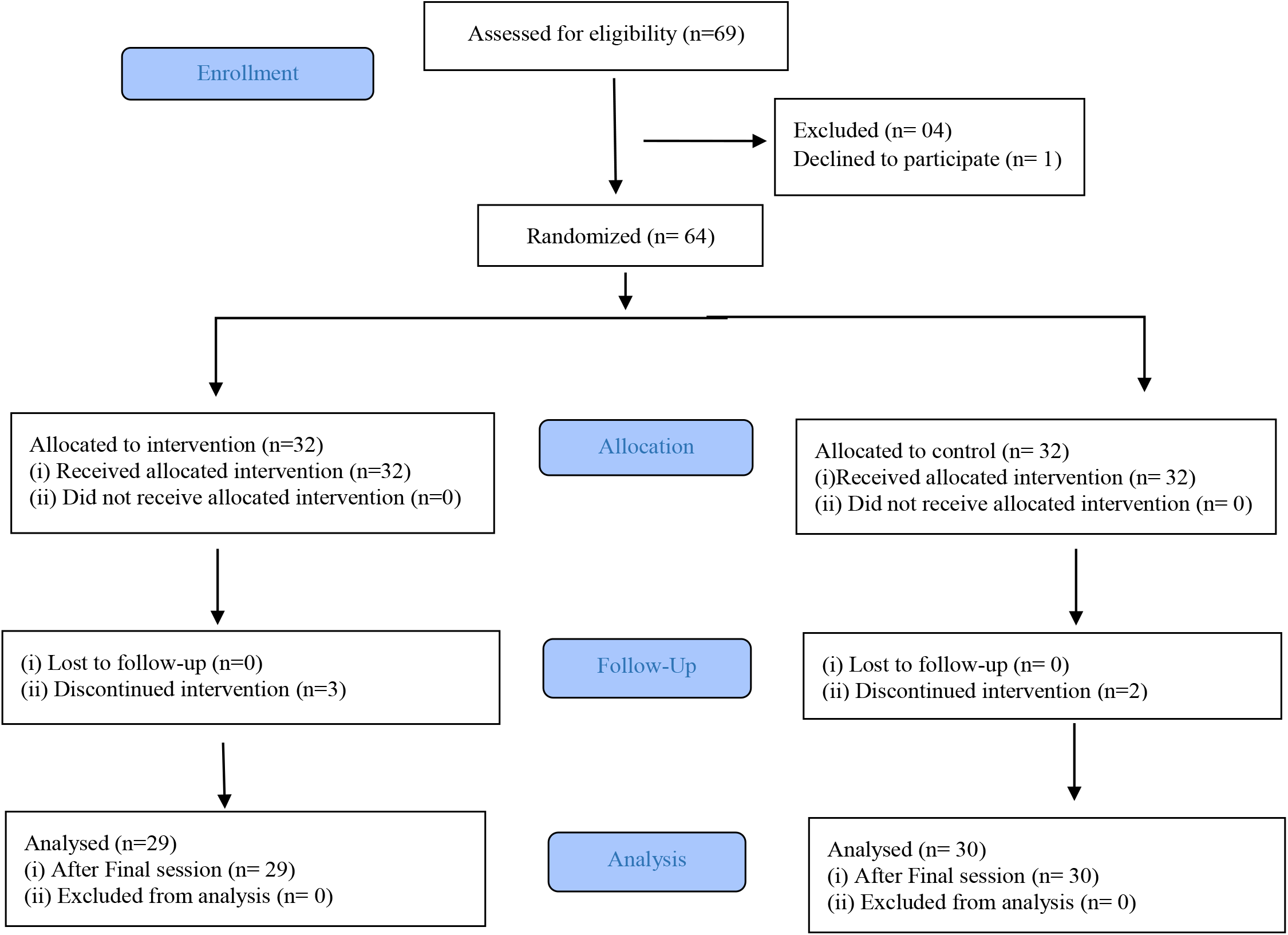
CONSORT 2010 flow diagram.

### Patients

From May 2020 to November 2021, 69 patients aged 30-60 years with a diagnosis of planter heel pain or plantar fasciitis or calcaneal spur by a physician according to ICD-10 [28] were recruited to the study through hospital randomization. Sixty-four (n=64) subjects complied with the eligibility criteria and were assigned, after voluntary written consent, to either SDM group or MFR by computer generated concealed allocation. The inclusion criteria were (i) subjects with diagnosis of unilateral or bilateral plantar fasciitis, heel spur, or plantar heel pain, according to the ICD 10 criteria (ii) Limited ankle dorsiflexion range of motion (ROM) in any range (iii) and pain more than 4 weeks. The exclusion criteria were (i) any history of fracture of foot or lower tibia in last 6 weeks, (ii) co-morbidity associated with infectious condition of foot, endocrine disease with visible cyanotic symptoms in foot, severe osteopenia of foot in x-ray or carcinoma, (iii) pre-existing phobia to physiotherapy or manipulative therapy. Both groups received interventions from two outpatient settings of a hospital. Interventions were given by an expert physiotherapist with extensive in-service training to follow the specific treatment protocol. The single assessor was blinded to the assignment and performed all the assessments. Baseline data were collected before treatment and repeated after 12 sessions (3 sessions, 4 weeks) of treatment in the hospital setting.

### Interventions

The MFR group received myofascial release of plantar fascia and perifascial structures in supine position [21]. The maneuver was explained and the subjects received three exercise interventions for 5-7 repetitions, with 15-30 seconds hold in progressive manner self-performed by subjects aided by the physiotherapist. The SDM group also received three exercise interventions from category A, B. C in a progressive manner to gastrocnemii and soleus muscles (figure 3) in same dosage, Duration and application process mostly performed by physiotherapist. In addition, both groups received strengthening exercise for global mobilizer of ankle and other conventional treatment such as Ultrasound therapy, shoe modification advice, ice or hot compression as advised by physiotherapist. From the day of baseline assessment, each participant received a total of 30 minutes of interventions, 3 times a week for 4 weeks.

**Figure 3:**
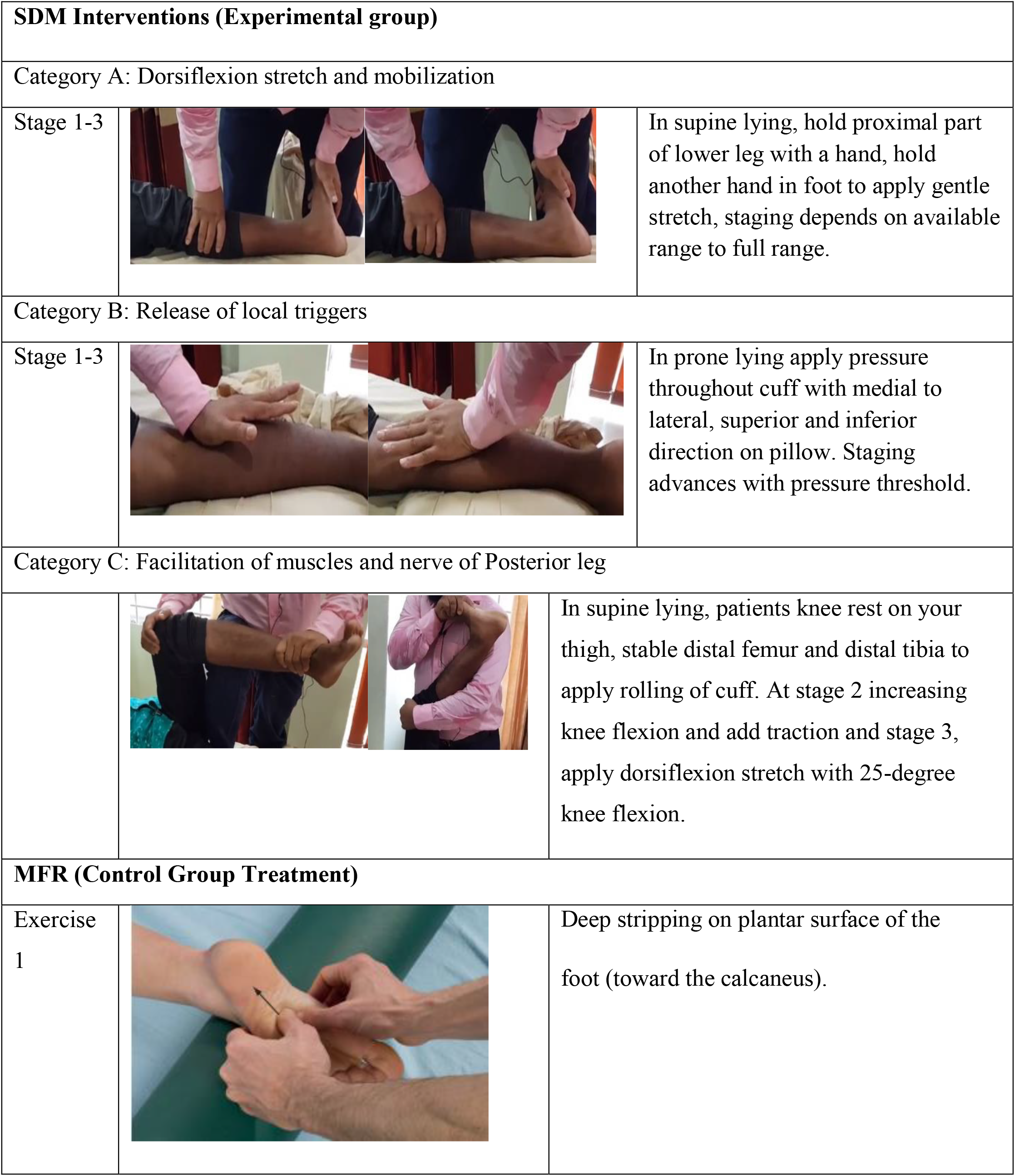
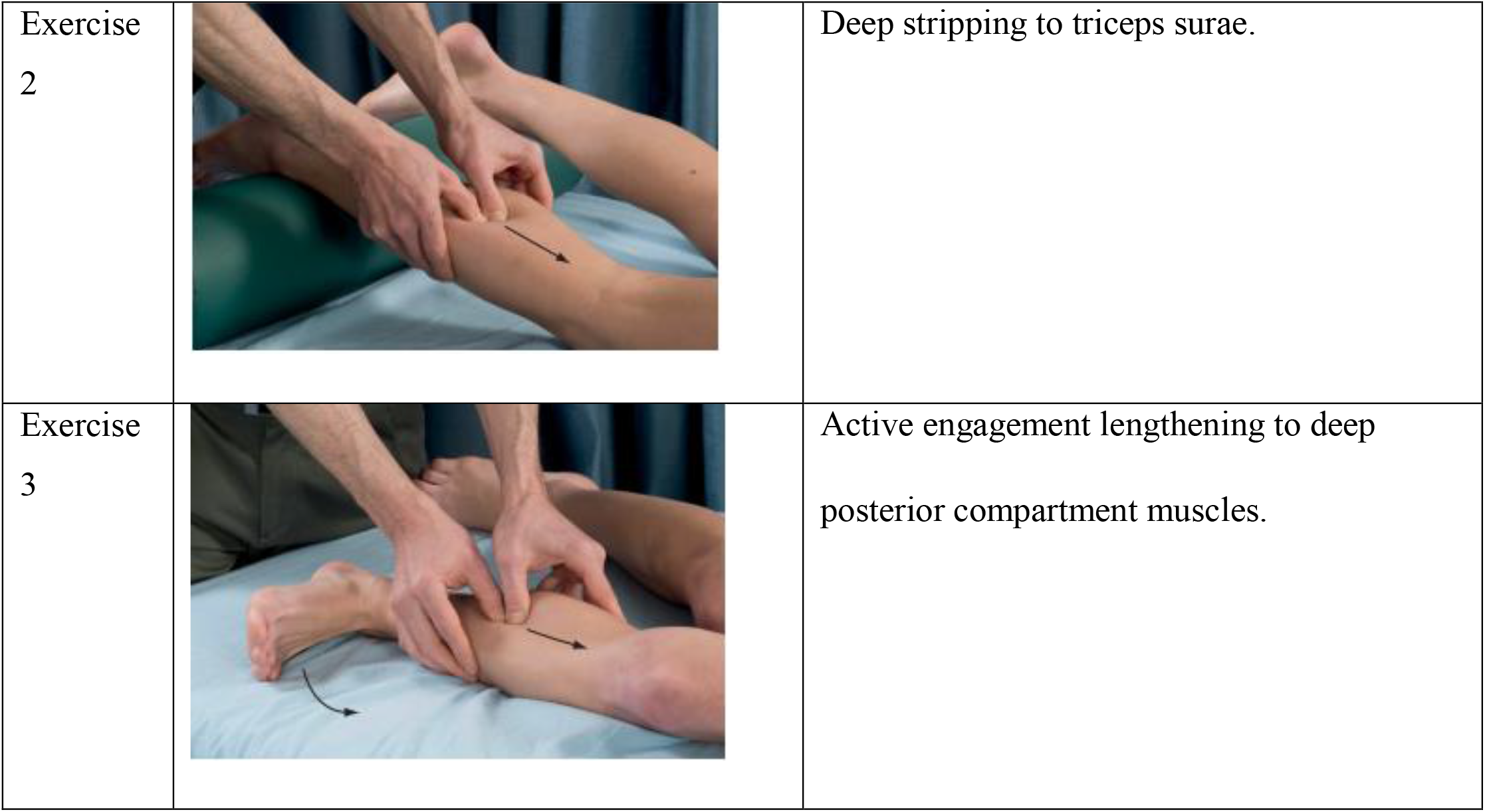
Interventions.

### Outcome measurement

Primary outcomes such as, pain, activity limitations and disability due to pain were assessed by foot function index (FFI) and range of motion of ankle dorsiflexion and plantar flexion by universal goniometer. Secondary outcomes evaluated were overall disability by foot ankle disability index (FADI) and ankle dorsiflexion and plantar flexion strength by 10-point manual muscle testing process. To ensure reliability an assessor blinded to the randomization assessed the baseline and post-test data with the aid of an independent data collector.

### Statistical analysis

Data entry and checking the quality of data were examined by a separate operator, blinded to the data collection process. Data were analyzed using a general linear model and input to SPSS Version 20.0. ROM, FADI and MMT were analyzed using parametric, paired t-test and independent t-test was used to compare baseline data to post-intervention data, with alpha set at .05. The chi-square test and independent-samples t-test were used to compare and analyze the clinical baseline characteristics between the groups.

## Results

Baseline variables were reported for the patients completing the intervention protocol (control 30, SDM 29) (table 3). There was statistically significant (<.05) association in age (p=.007), BMI (p=.001), occupation (p=.009), working habit (p=.001), Duration of symptom (p=.046) and disability in FADI (p=.001) found in baseline between groups.

**Table 3:** Baseline variables of all subjects.

| Variable | MFR (n=31) | SDM (n=29) | P |
| --- | --- | --- | --- |
| Age (years) | 42.5 ± 8.7 | 48.2 ± 6.7 | .007 <sup>1</sup> |
| Gender (M/F) | 9/22 | 7/22 | .668 <sup>2</sup> |
| BMI | 25.7 ± 2.2 | 30.8 ± 4.8 | .001 <sup>1</sup> |
| Occupation |  |  |  |
| Farmer | 1 | 3 | .009 <sup>2</sup> |
| Garments Worker | 6 | 2 |  |
| Businessman | 4 | 0 |  |
| Housewife | 16 | 20 |  |
| Teacher | 4 | 0 |  |
| Technical Professionals | 0 | 4 |  |
| Walking habit daytime |  |  |  |
| Sandal | 9 | 7 | .001 <sup>2</sup> |
| High Heel | 12 | 2 |  |
| Shoe | 8 | 4 |  |
| Walk Barefoot | 2 | 16 |  |
| Working hours | 7.1 ± 1.7 | 7.5 ± 1.1 | .308 <sup>1</sup> |
| Limb affected (1/2) | 28/3 | 26/3 | .931 <sup>2</sup> |
| Duration (Weeks) | 11.2 ± 11.2 | 19.0 ± 17.8 | .046 <sup>1</sup> |
| AROM PF | 38.2 ± 3.5 | 38.4 ± 3.5 | .809 <sup>1</sup> |
| AROM DF | 27.4 ± 2.8 | 28.5 ± 2.7 | .123 <sup>1</sup> |
| Strength PF (median, mode) | 7/7 | 6/6 | .122 <sup>2</sup> |
| Strength DF(median, mode) | 6/6 | 7/6 | .509 <sup>2</sup> |
| Pre FFI Pain | 60.4 ± 4.7 | 60.2 ± 7.9 | .885 <sup>1</sup> |
| Pre FFI disability | 60.8 ± 5.0 | 62.5 ± 10.1 | .426 <sup>1</sup> |
| Pre FFI activity | 30.2 ± 6.8 | 24.4 ± 8.7 | .006 <sup>1</sup> |
| Pre FADI total | 70.6 ± 8.3 | 78.7 ± 7.3 | .001 <sup>1</sup> |

<sup>1</sup>Independent sample t test, <sup>2</sup>Chi square test, significance level (<.05)
MFR: Myofascial Release
SDM: Structural Diagnosis and Management

The respondents were mostly in their 4^th^ decade of life (control 42.5 ± 8.7, SDM 48.2 ± 6.7), female majority with overweight (BMI control 25.7 ± 2.2, SDM 30.8 ± 4.8), housewife (n=36) working nearly 8 hours a day and with chronicity (Control 11.2 ± 11.2 weeks, SDM 19.0 ± 17.8). In both groups dorsiflexion range was limited nearly half of the range (Control 27.4 ± 2.8 degrees and SDM 28.5 ± 2.7 degrees).

Both MFR and SDM group had statistically significant improvements from baseline in all the variables. In paired sample t test, MFR group had mean, lower and upper margin of 95% CI as FFI pain (30.38, 27.9, 32.8, p=.001), FFI disability (31.29, 28.64, 33.93, p=.002), FFI activity (14.77, 12.71, 16.83, p=.001), FADI (−21.03, −24.86, −17.20, p=.04), Plantar flexion range (−7.742, −11.40, −4.079, p=.014) and dorsiflexion range (−10.96, −12.76, −9.170, p=.02); SDM group had mean, lower and upper margin of an 95% CI was as FFI pain (29.73, 23.96, 35.62, p=.001), FFI disability (29.12, 23.58, 34.67, p=.001), FFI activity (10.35, 9.114, 11.59, p=.001), FADI (−7.677, −11.48, −3.870, p=.001), Plantar flexion range (−5.806, −10.25, −1.362, p=.012) and dorsiflexion range (−6.677, −8.404, −4.951, p=.001). In Wilcoxon test, the MFR group had Z value, alpha value and lower and upper value related to the alpha value was strength in plantar flexion (−3.93, .003, .001, .095) and in dorsiflexion strength (−4.89, .002, .001, .095); subsequently in SDM the value was PF (−4.544, .003, .001, .095) and (−4.763, .002, .001, .095).

In between group analysis (table 2), SDM was found to be superior than MFR with statistical significance in FFI pain (p=.001), FFI activity (p=.009), FFI disability (p= .001) and FADI (p=.002) in independent sample t test; also, Mann Whitney U test analysis found plantar flexion strength to be significant in SDM group (P=.033), other parameters found clinical significance rather than statistical difference. Detailed results are appended in table 4.

**Table 4:** Group comparison using paired and independent sample t test and Willcoxon and Mannwhitney U Test.

| Variables | MFR |  |  | SDM |  |  | Between group |  |  |  |
| --- | --- | --- | --- | --- | --- | --- | --- | --- | --- | --- |
|  | mean | 95% CI | p | mean | 95% CI | p | md | t | SED | p |
| **FFI Pain** | 30.38 | 27.9- 32.8 | .001 | 29.73 | 23.96- 35.62 | .001 | -.349 | -.221 | 1.578 | .001 |
| **FFI Disability** | 31.29 | 28.64- 33.93 | .002 | 29.12 | 23.58- 34.67 | .001 | -<br>3.075 | -2.28 | 1.348 | .001 |
| **FFI Activity** | 14.77 | 12.71-16.83 | .001 | 10.35 | 9.114- 11.59 | .001 | 1.277 | .707 | 1.807 | .009 |
| **FADI** | -<br>21.03 | -24.86, -<br>17.20 | .04 | -<br>7.677 | -11.48, -<br>3.870 | .001 | 5.261 | 3.531 | 1.807 | .002 |
| **ROM PF** | -<br>7.742 | -11.40, -<br>4.079 | .014 | -<br>5.806 | -10.25, -<br>1.362 | .012 | 2.002 | .719 | 2.002 | .176 |
| **ROM DF** | -<br>10.96 | -12.76, -<br>9.170 | .02 | -<br>6.677 | -8.404, -<br>4.951 | .001 | 3.215 | 3.890 | .826 | .456 |
|  | Z | p | 95% CI | Z | p | 95% CI | U | p | 95% CI |  |
| **Strength PF** | -3.93 | .003 | .001- .095 | -<br>4.544 | .003 | .001-.095 | 305 | .033 | .001-.079 |  |
| **Strength DF** | -4.89 | .002 | .001- .095 | -<br>4.763 | .002 | .001-.095 | 348 | .150 | .060-.240 |  |

Significance level (<.05)

## Discussion

The randomized clinical study found SDM is effective to reduce pain, improve activities, ankle range of motion ROM, and remission of disability in plantar heel pain. Moreover, SDM approach is superior in remission of pain, improving activity and reducing disability for the participants with plantar heel pain. The respondents were mostly in the age range 30-40 years, female, overweight and working for more than 8 hours a day. They also had limited dorsiflexion range nearly half of the full range. Sullivan and colleagues [2] found that the plantar heel pain is associated with a higher BMI, decreased ankle dorsiflexion range of motion and reductions in some extent of foot and ankle strength and flexibility. The mentioned study also found no association with plantar fascia flexibility, nor changes of arch with plantar heel pain. This could justify stretching the gastrocnemii and soleus muscles progressively, rather than stretching the plantar fascia and gastrocnemii and soleus muscles, collectively. Although DiGiovanni and colleagues found the opposite result, it was the process of stretching that made the difference; the study instructed the patient to stretch the calf (gastrocnemii and soleus muscles) in a standing position, keeping the affected limb behind contralateral limb in a straight-line, while keeping the knee extended [20]. Garten [29] explained a different process of stretching of the calf in myofascial pain syndrome. He described dorsiflexion stretch keeping knee extended only stretches the gastrocnemii, to stretch soleus the knee should be flexed. The SDM approach integrates stretching to the gastrocnemii (A), myofascial release (B) and stretch to the soleus muscles that follow the principles of structural correction. These structural correction measures are proven to improve ankle flexibility and correct the biomechanics of “windlass mechanism” of the foot [26]. This study found significant reduction in pain and disability and improvements inactivity, range for both groups, compared to baseline (table 4), These findings concur with previous findings and provide evidence that stretching to the plantar fascia, gastrocnemii and soleus muscles [20] or stretching gastrocnemii and soleus muscles, alone, have a significant (p=<.05) improvement effect, compared to baseline, in 4 weeks (12 sessions). Although the MFR process stated in the control was not supported by Garten [29], the reason of improvement might be the stated process of plantar fascia release. Between group analysis found the SDM intervention for releasing gastrocnemii and soleus muscles was significantly superior in reducing pain and disability and improving activity (p=<.005) compared to the MFR process [20] of gastrocnemii, soleus and plantar fascia. Thus, it can be concluded that the SDM intervention for gastrocnemii and soleus is superior to conventional MFR for gastrocnemii, soleus and plantar fascia. This indicates, there is no need for an intervention to plantar fascia (local structure) if MFR Intervention applied targeting the gastrocnemii and soleus. The study also found significant changes in dorsiflexion and plantar flexor strength in both groups, with both groups receiving conventional strengthening and other interventions, per treatment protocol. Future studies should include a repeated measures evaluation, to explore the long-term effects of the SDM intervention. Also, a larger sample size would allow application of parametric statistical analysis, measuring strength with a standardized test, such as a dynamometer, and inclusion of gait and posture analysis may provide greater understanding of the multi-dimensional context and outcomes for the SDM approach.

## Conclusion

MFR and SDM approaches are both effective interventions to reduce pain and disability, improve activities and ankle ROM, in patients with plantar heel pain. However, the SDM approach is the method of choice in reduction of pain and disability and for improving activity.

## Data Availability

All data produced in the present study are available upon reasonable request to the authors

## Notes

### Competing Interest Statement

The authors have declared no competing interest.

### Clinical Trial

CTRI/2020/05/025151

### Funding Statement

This study did not receive any funding

### Author Declarations

IRB of Bangladesh Health Professions Institute (BHPI) gave ethical approval for this work (CRP-BHPI/IRB/12/18/1285)

